## Supplemental Appendices for "What incentives encourage local communities to collect and upload mosquito sound data by using smartphones? A case study in the Democratic Republic of the Congo"

Storer *et al.*

The supplementary information contains the demographic questionnaire (*Supplement 1 Appendix,* pg. 2-15) and the English version of the focus group discussion questions (*Supplement 2 Appendix,* pgs. 16-18). These supplementary methods are followed by the supplementary results of demographic comparison between districts and experimental groups using Wilcoxon rank sum tests or Fisher’s Exact Test depending on sample group size and data type (*Supplement 3, Table,* pg. 19-20).

***S1 Appendix: Demographic Questionnaire***

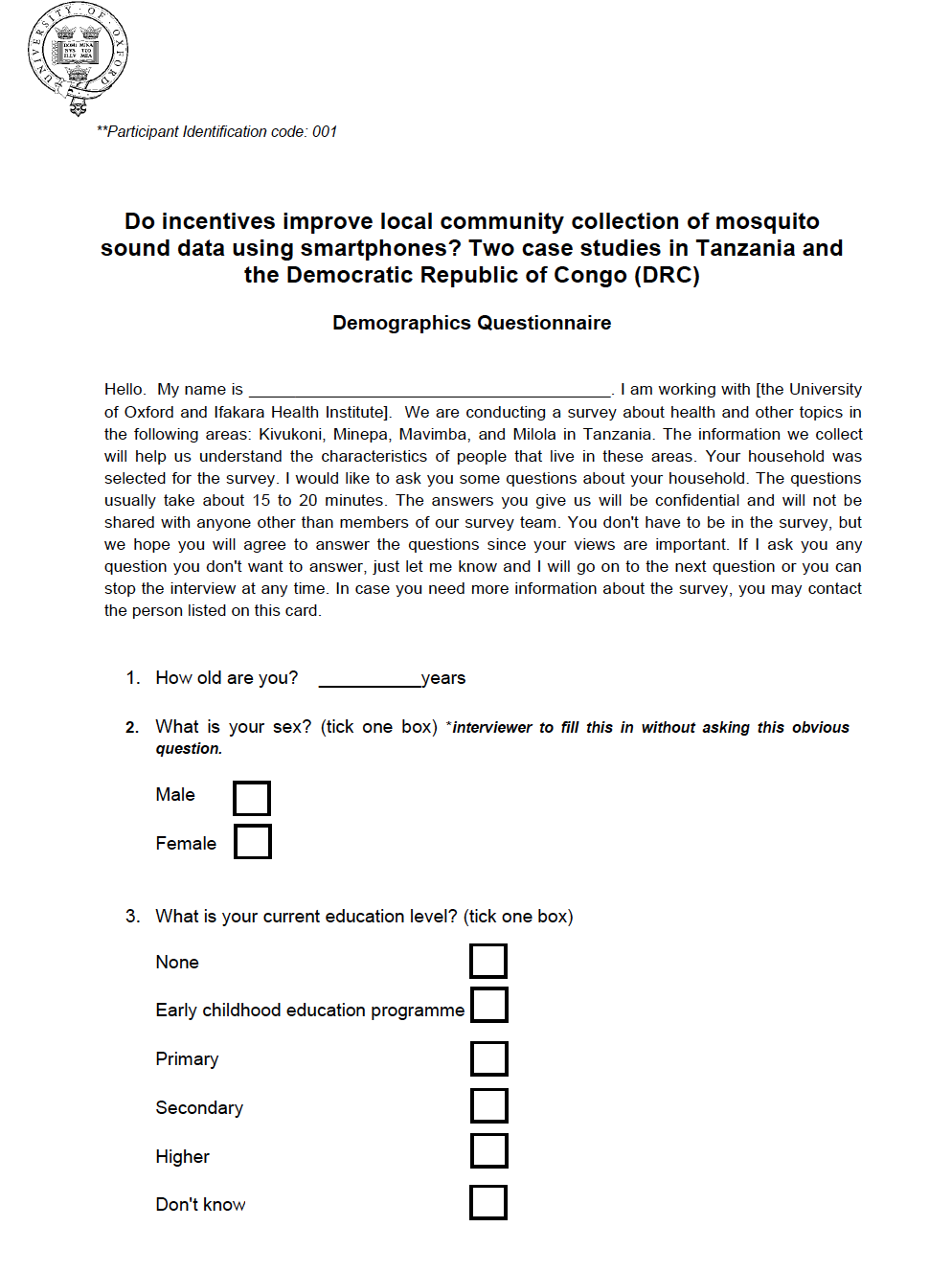

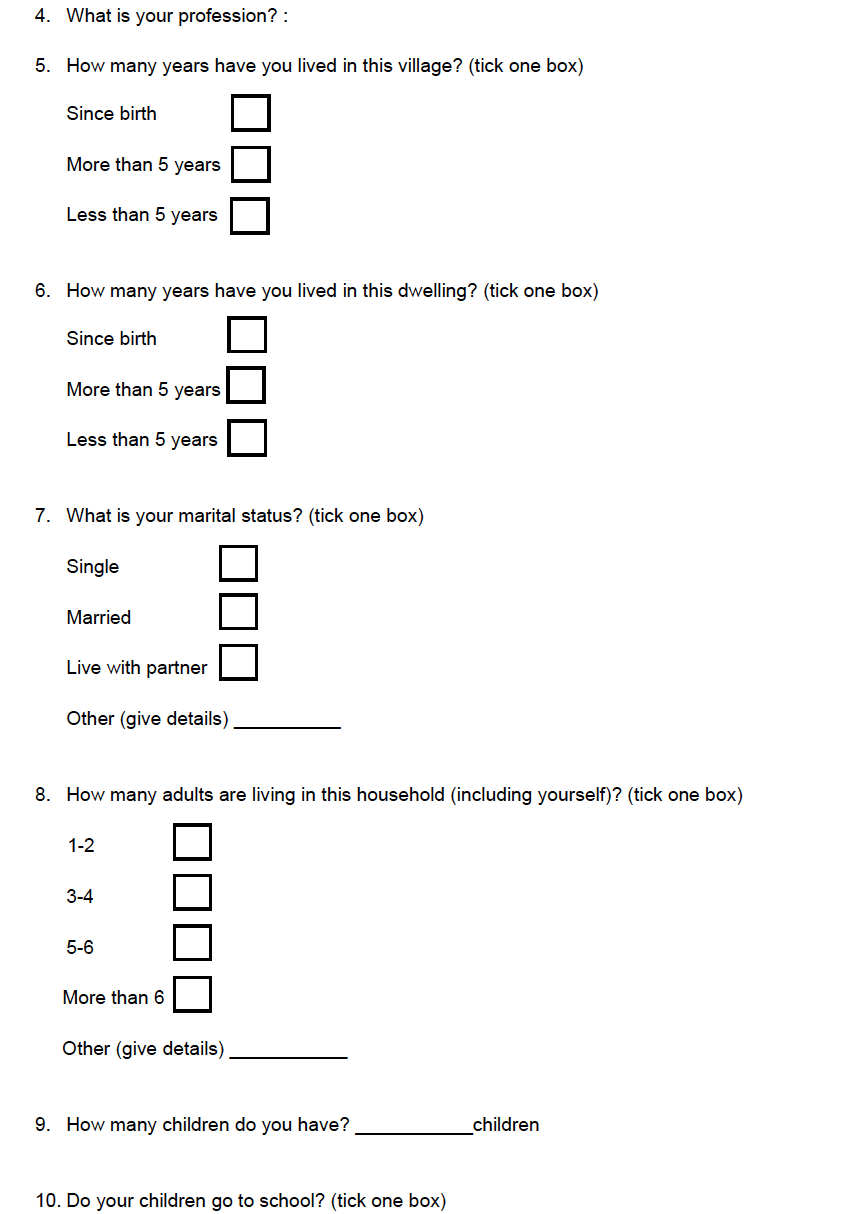

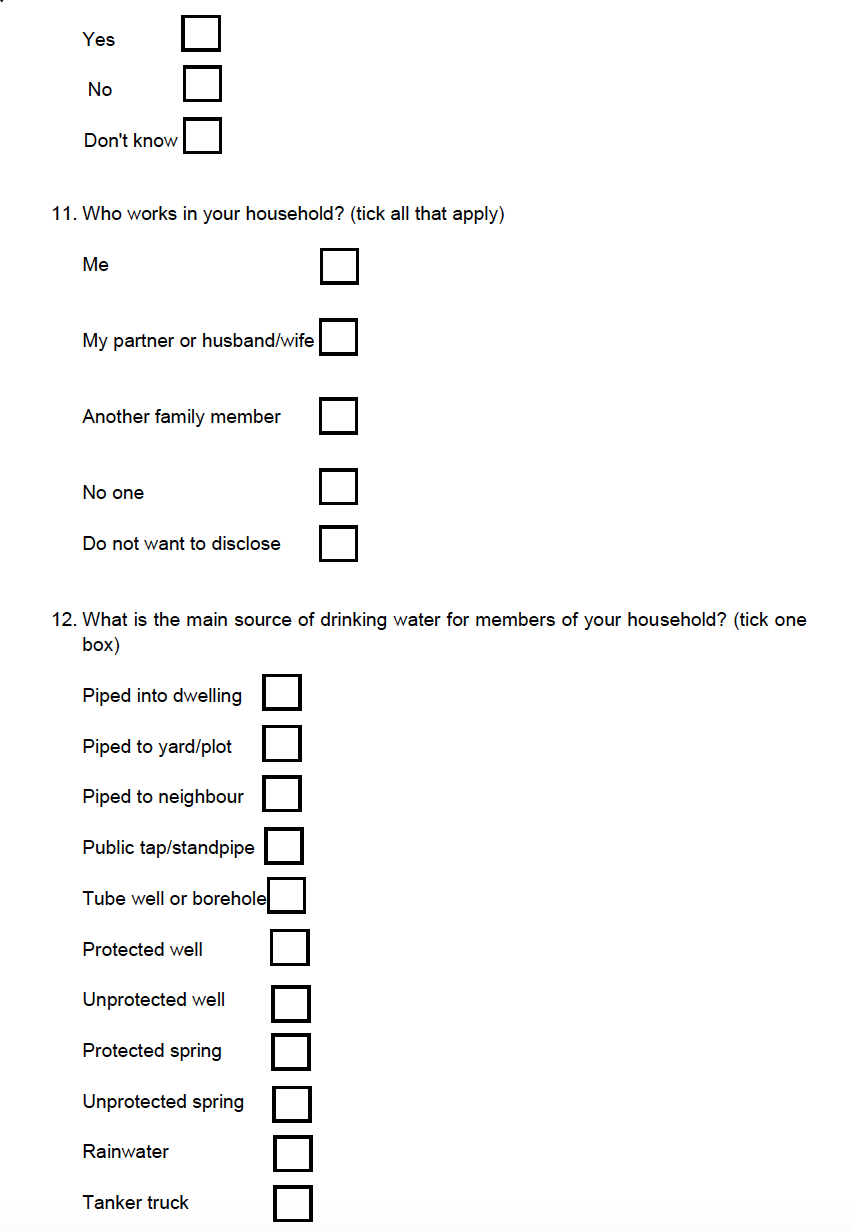

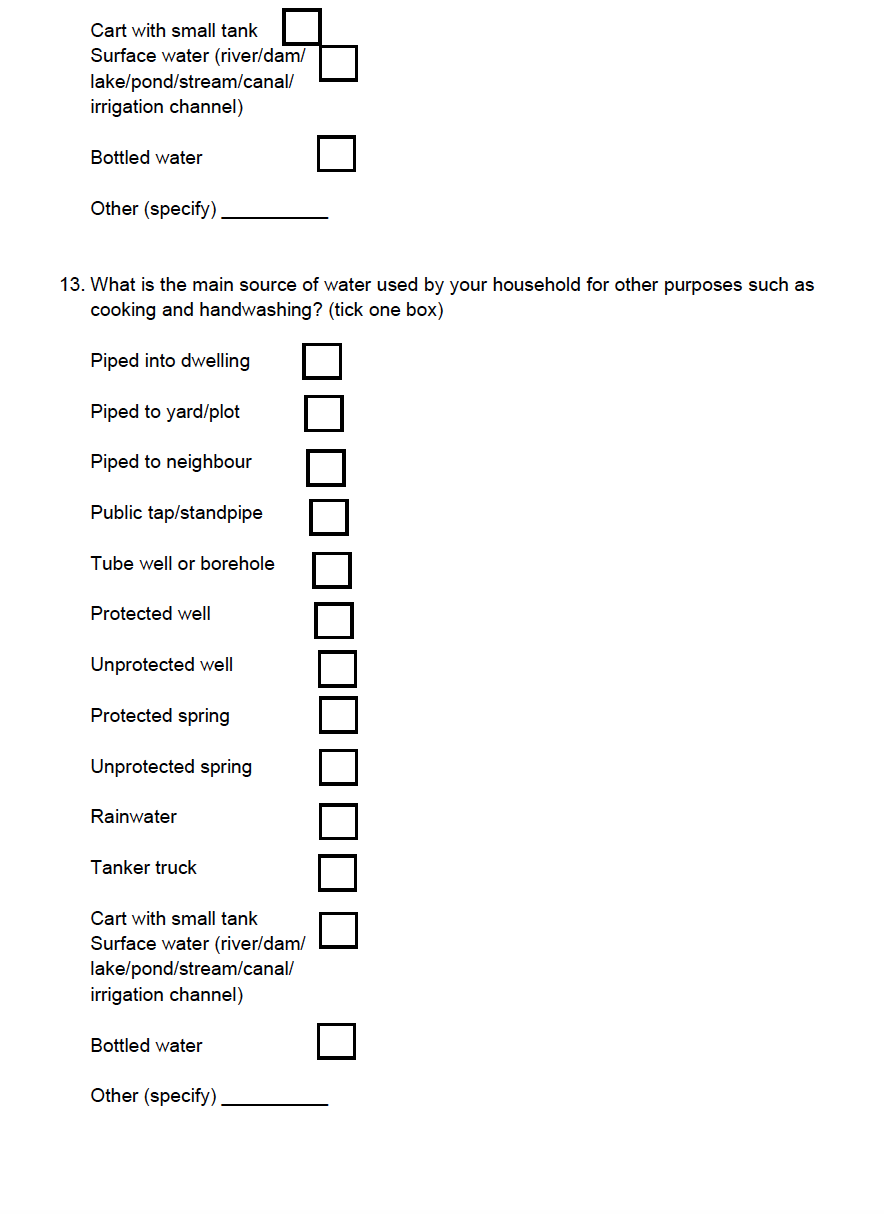

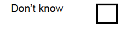

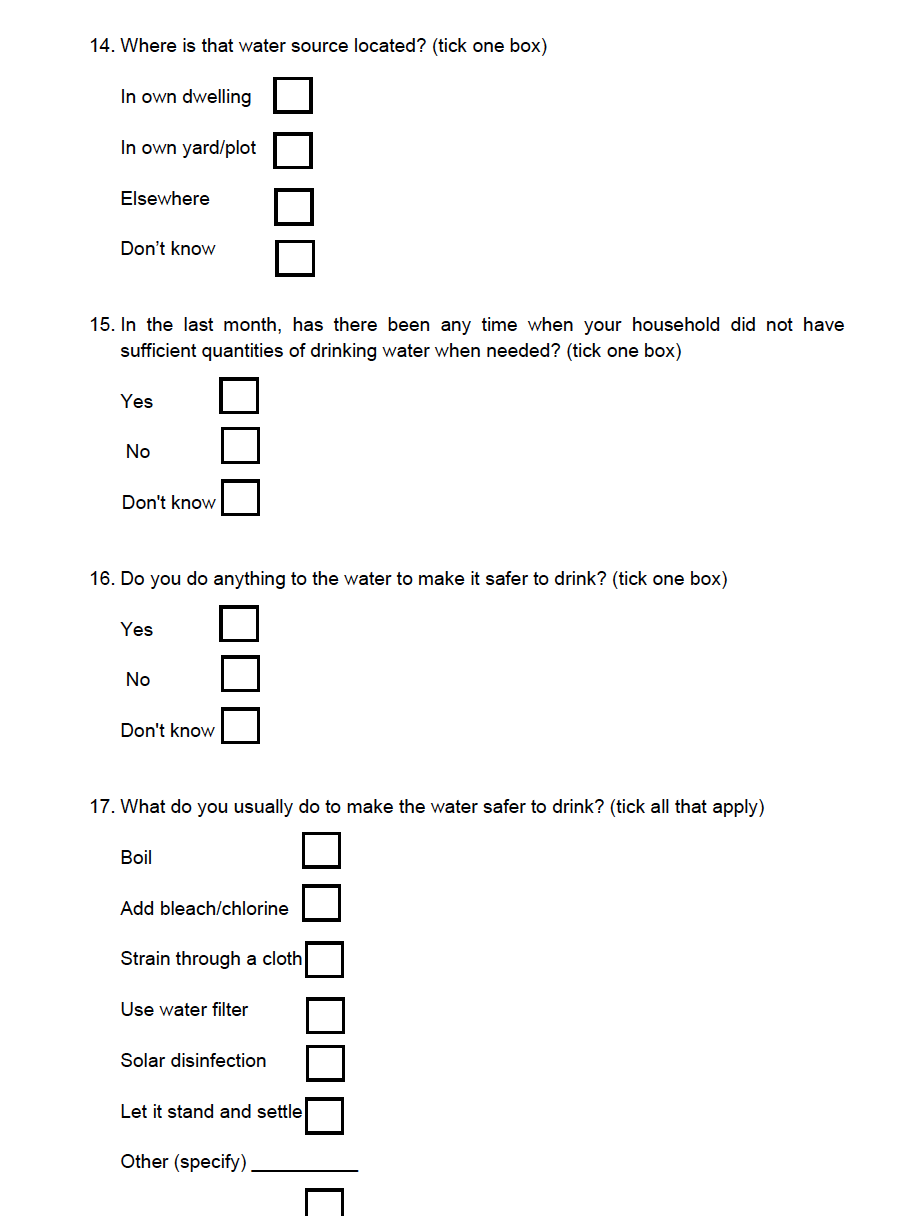

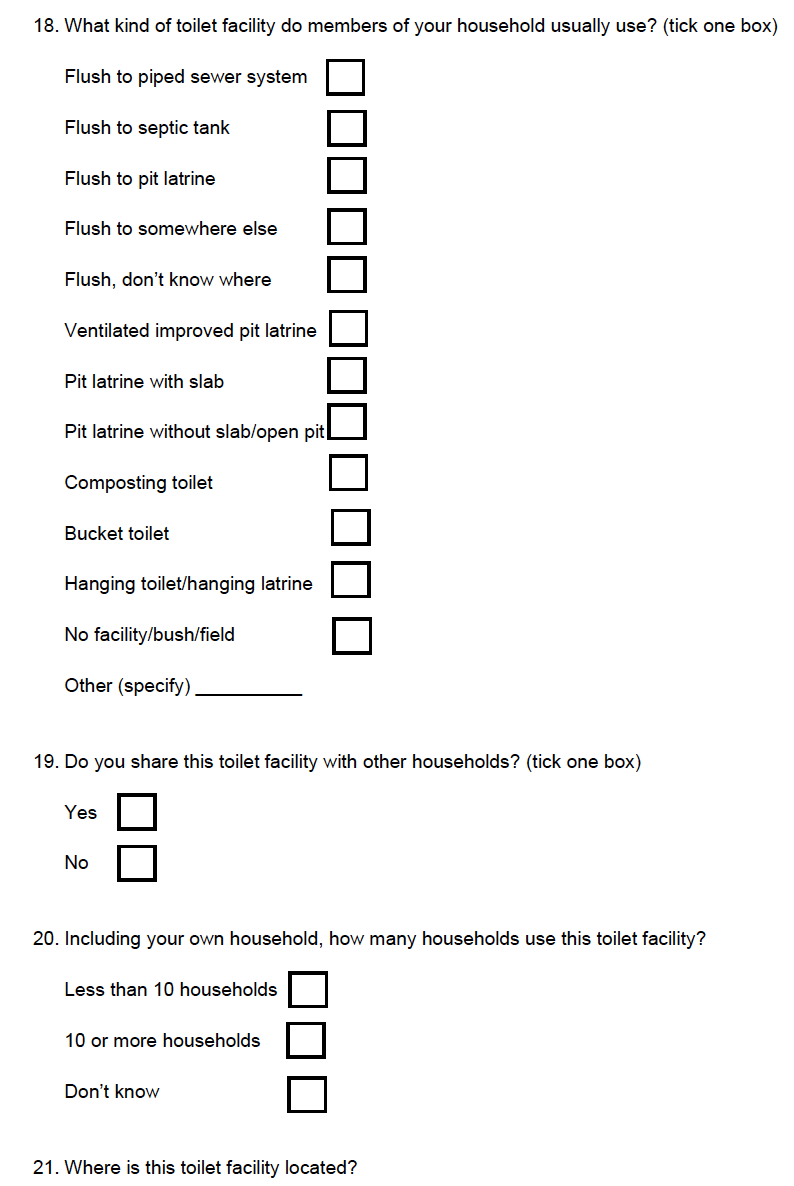

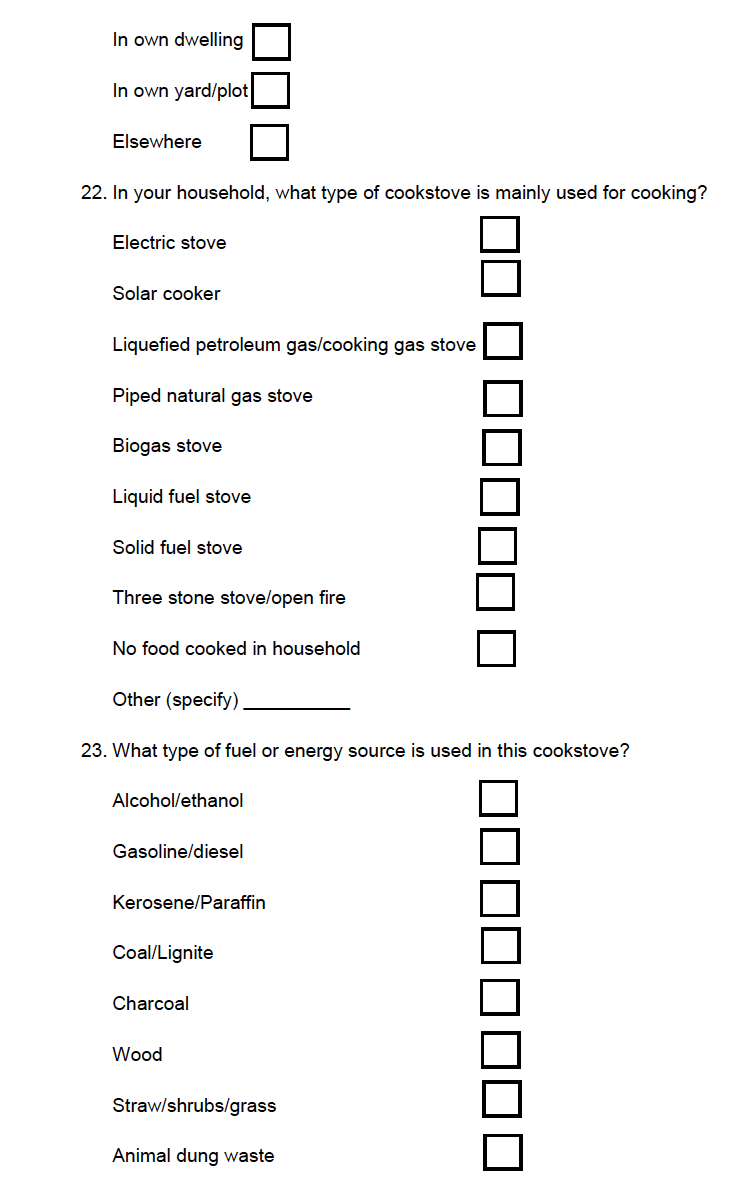

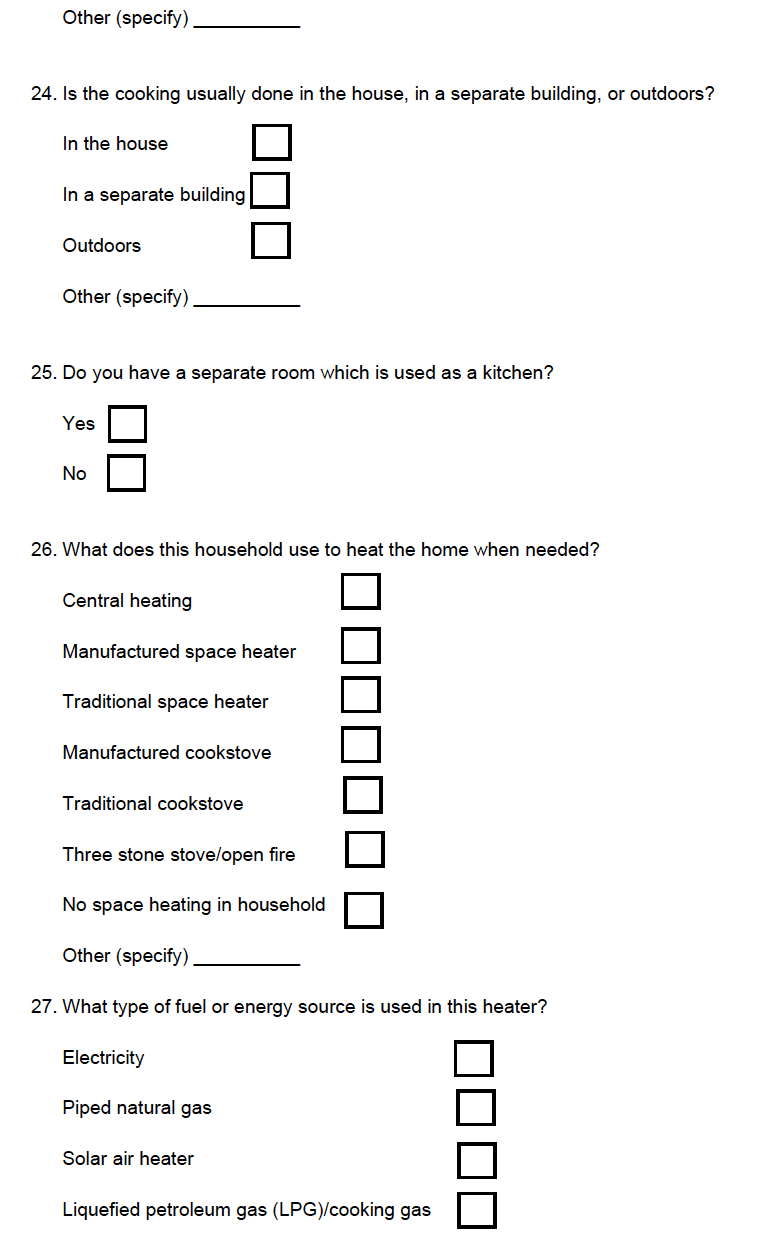

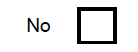

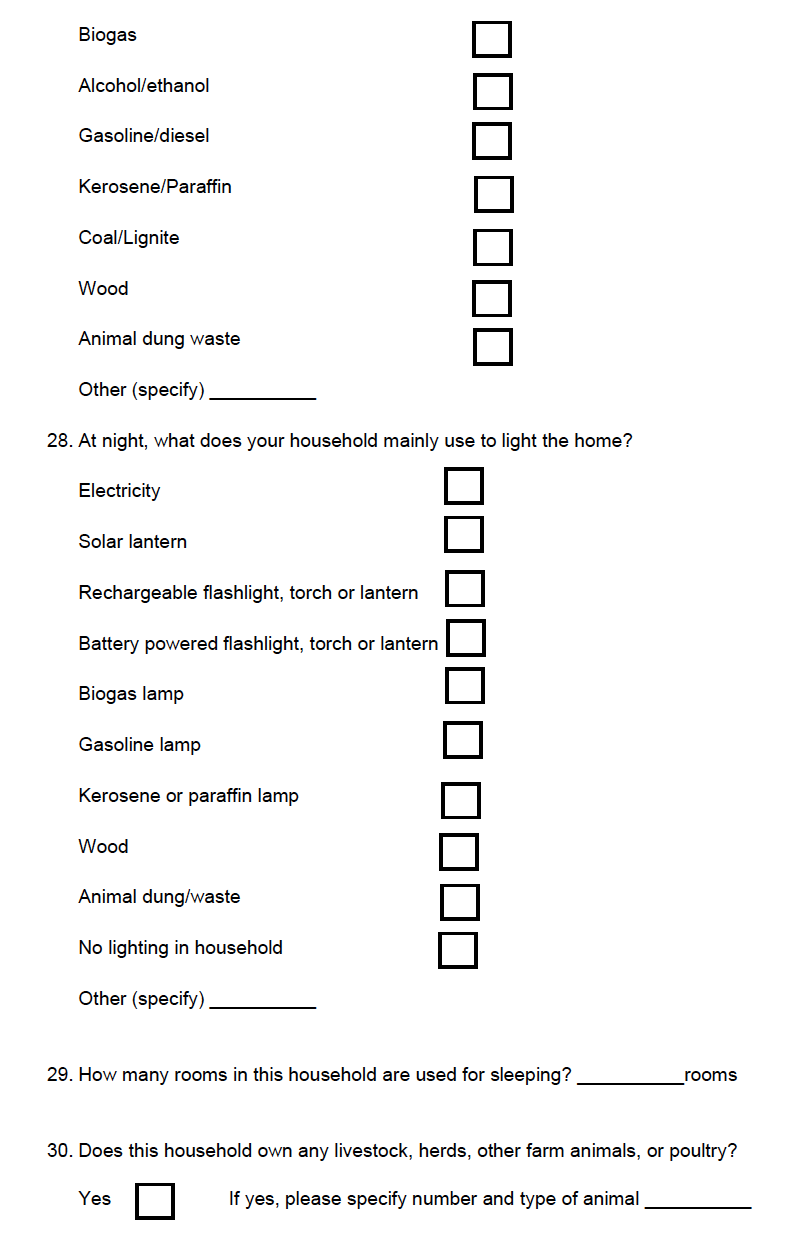

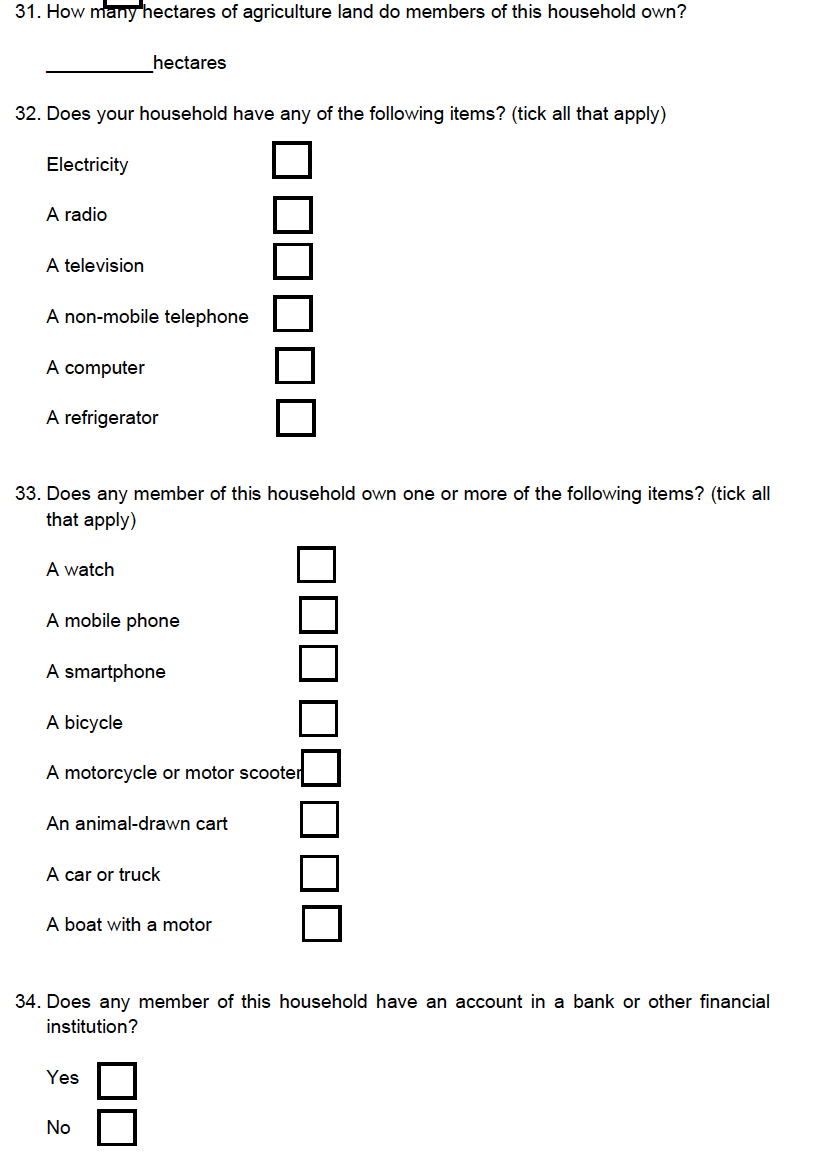

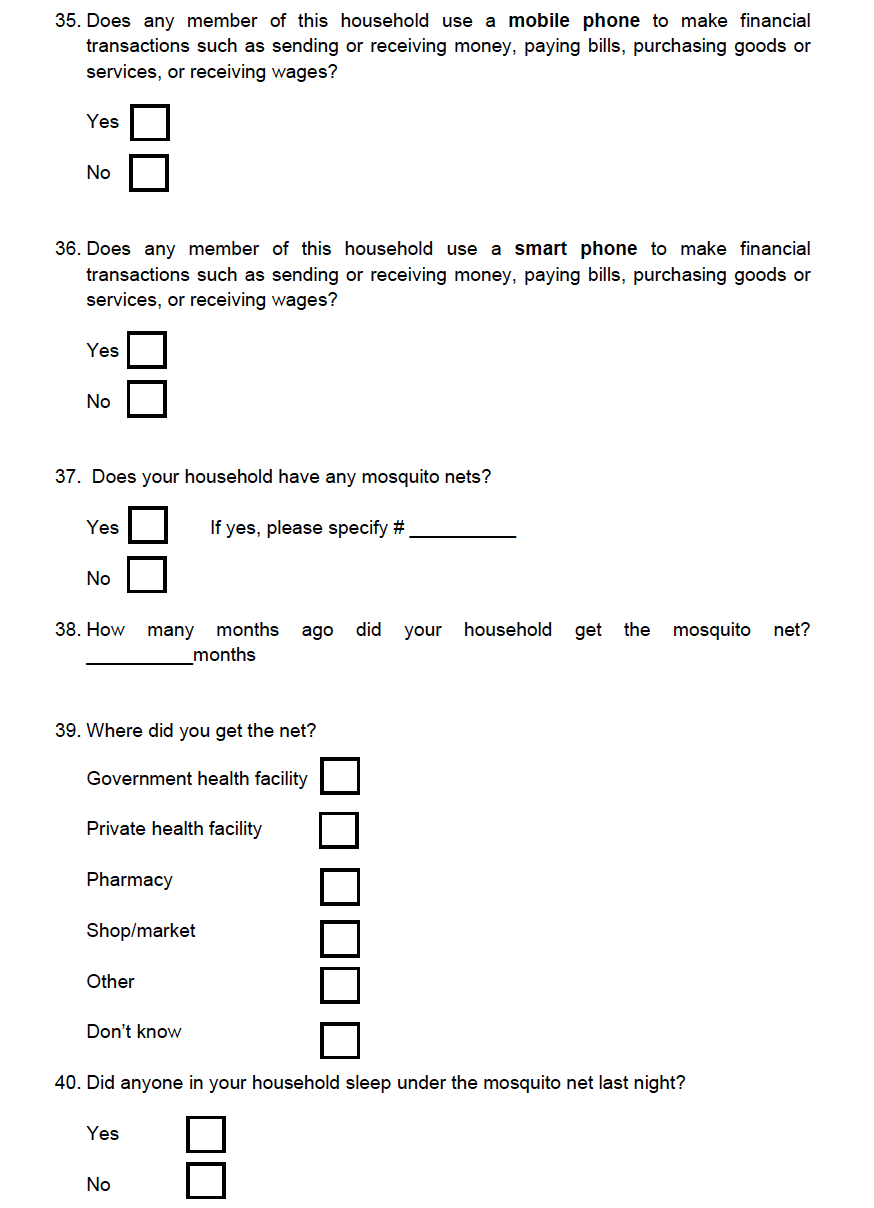

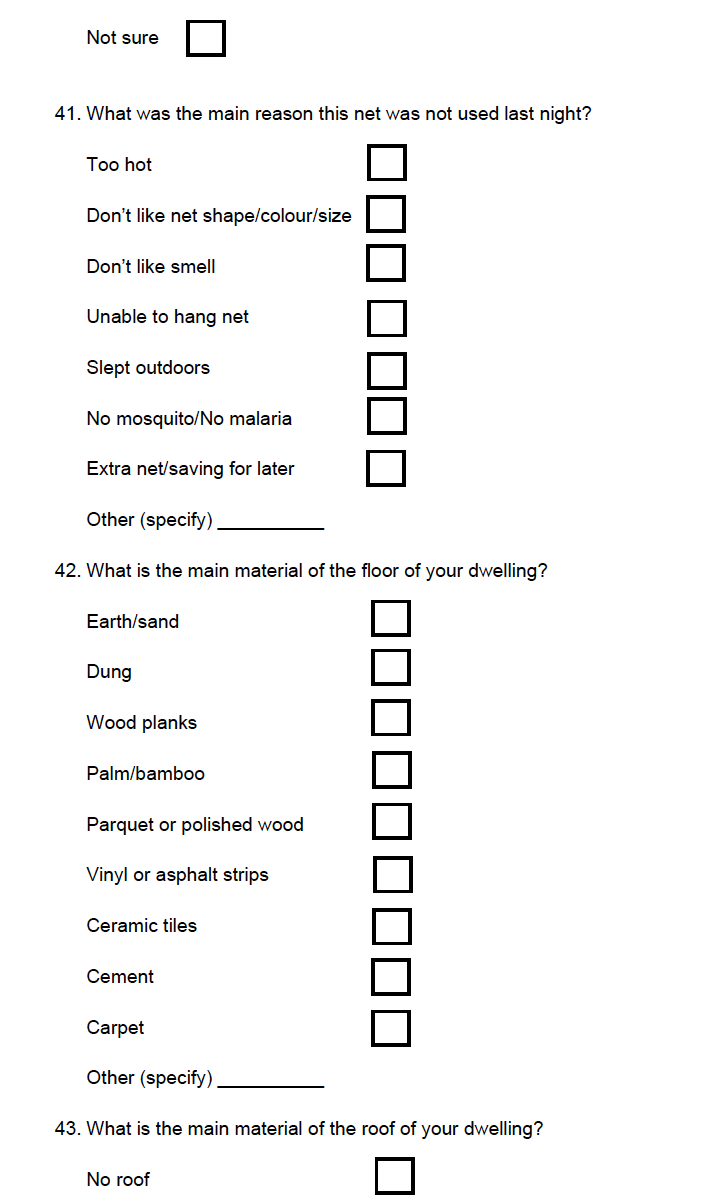

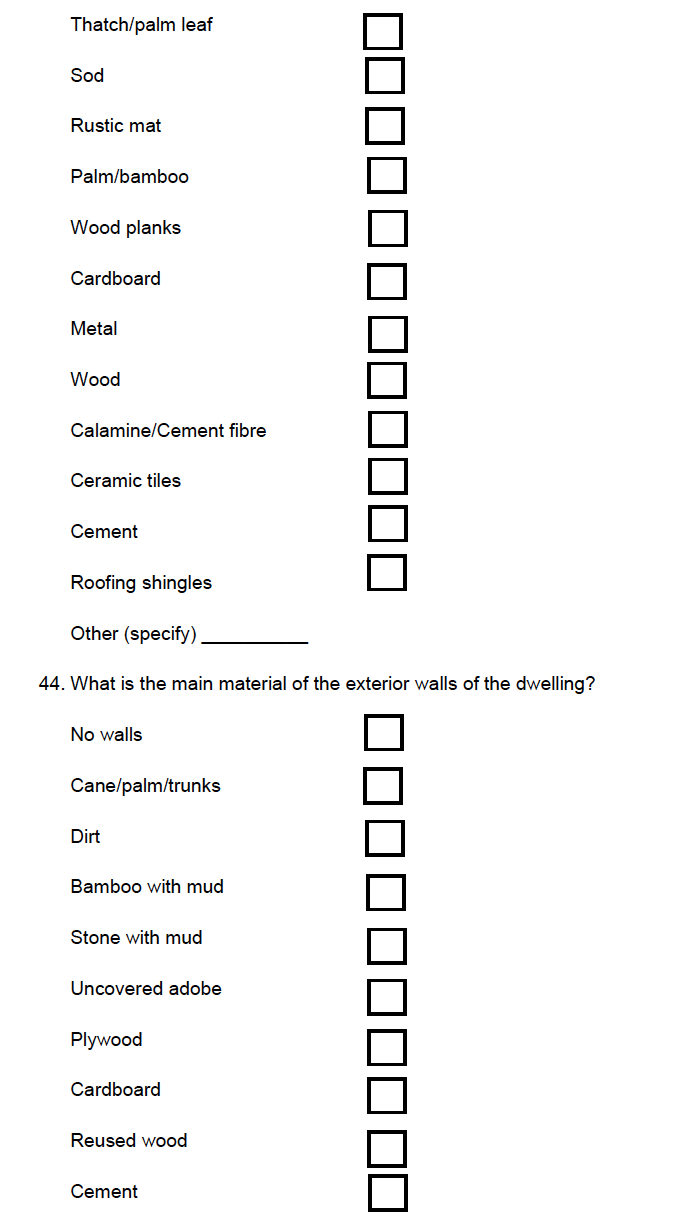

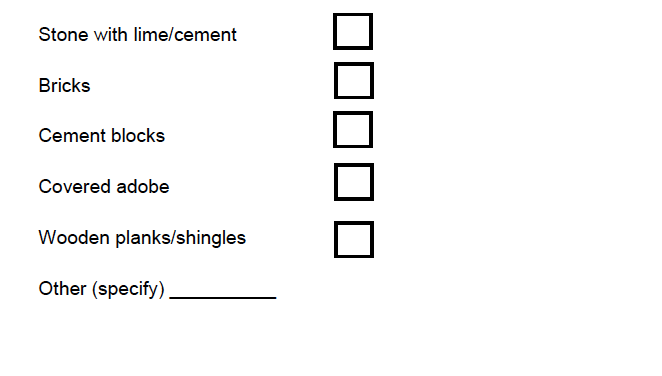

***S2 Appendix: Focus Group Discussions: Pre and Post Trial***

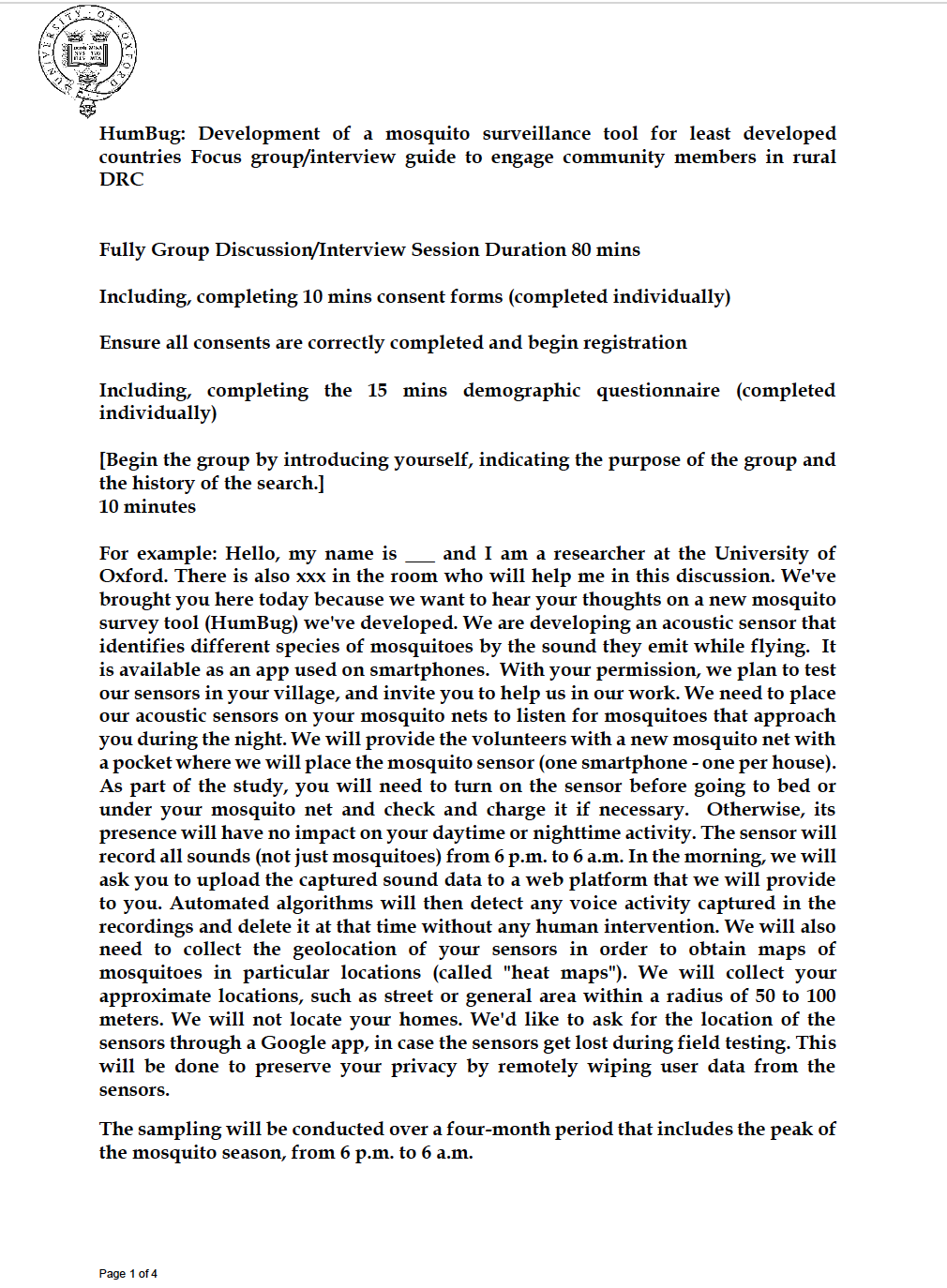

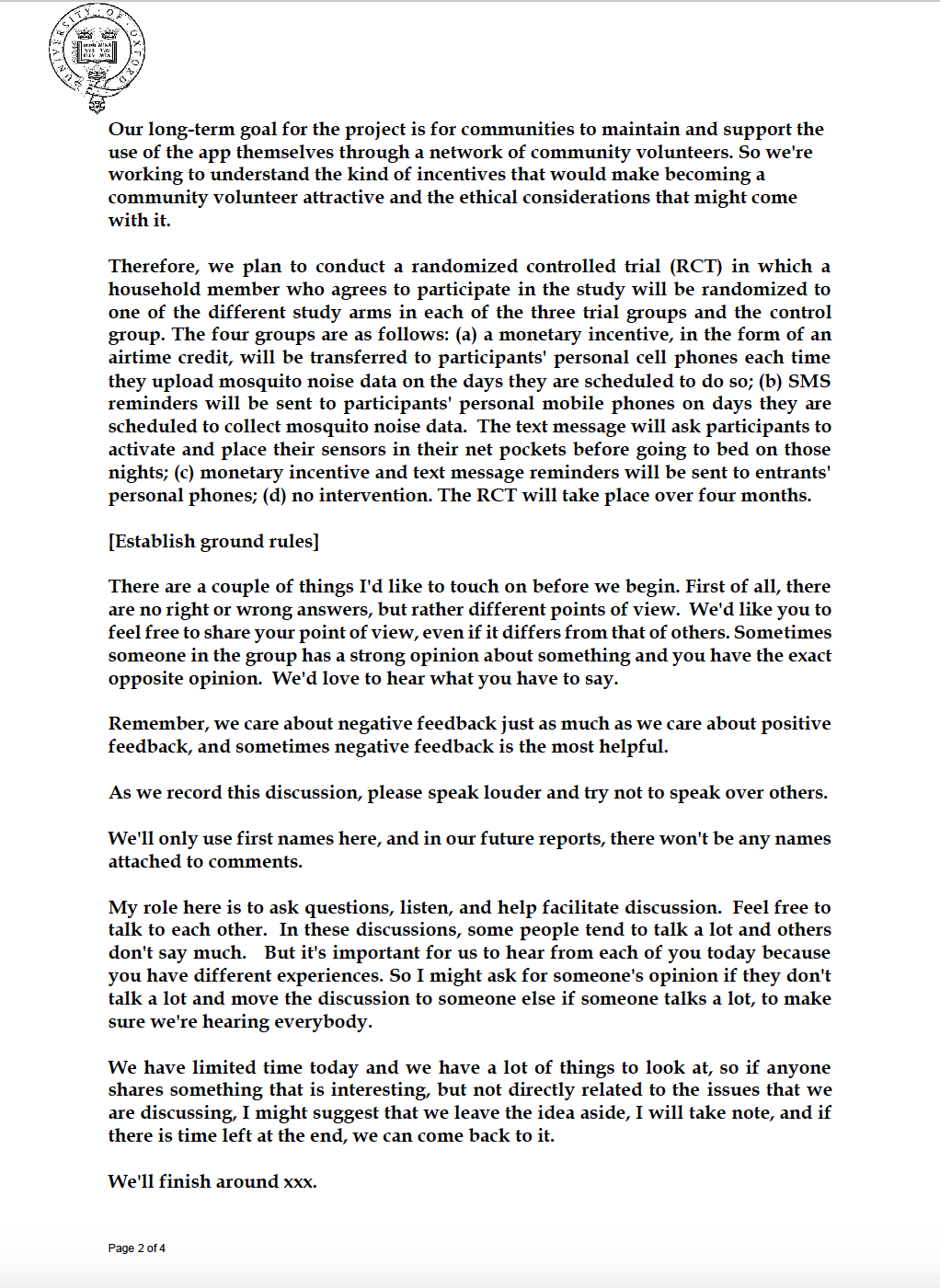

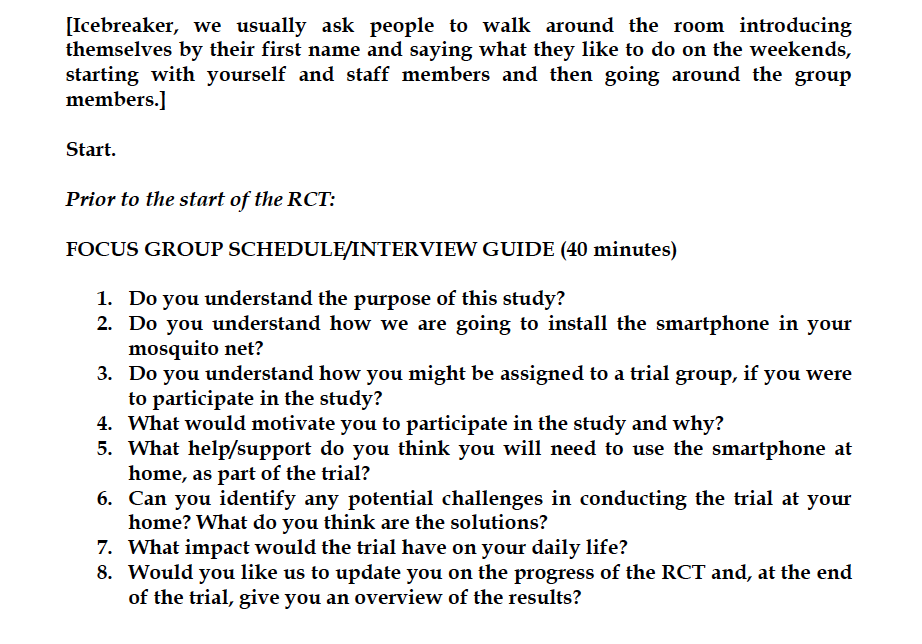

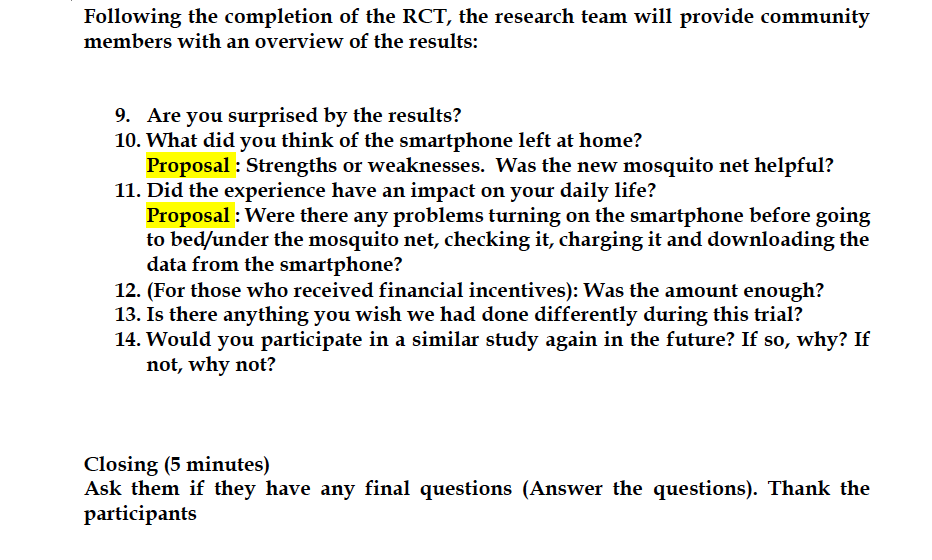

***Supplement 3: Table of Demographic Comparisons***

| Compared Groups | | | | | |
| --- | --- | --- | --- | --- | --- |
| Test Type | Compared Trait | Bandundu and Kinshasa | Control and Incentive | Kinshasa | Bandundu |
|  |  |  |  | Control and Incentive | Control and Incentive |
| Wilcoxon rank sum test with continuity correction | Age | W = 2383, p-value = 0.35 | W = 1843, p-value = 0.27 | W = 381, p-value = 0.16 | W = 428, p-value = 0.48 |
|  |  | 95% CI [-2, 7] | 95% CI [-7, 2] | 95% CI [-11, 2] | 95% CI [-9, 4] |
|  | Income | W = 2186.5, p-value = 0.32 | W = 1722.5, p-value = 0.41 | W = 299.5, p-value = 0.061 | W = 446, p-value = 0.98 |
|  |  | 95% CI [-20000, 80000] | 95% CI [-90000, 20000] | 95% CI [-1.70e+05, 5.15e-11] | 95% CI [-100000, 70000] |
|  | Years in Home | W = 2213.5, p-value = 0.84 | W = 2230.5, p-value = 0.34 | W = 480.5, p-value = 1 | W = 538, p-value = 0.31 |
|  |  | 95% CI [-1.88e-11, 1.04e-11] | 95% CI [-2.26e-12, 3.80e-11] | 95% CI [-2.38e-12, 2.38e-12] | 95% CI [-7.94e-11, 8.65e-12] |
|  | Number of Adults in Home | W = 1680, p-value = 0.017 | W = 2110, p-value = 0.87 | W = 510, p-value = 0.67 | W = 517.5, p-value = 0.55 |
|  |  | 95% CI [-1.00e+00,-3.50e-11] | 95% CI [-6.75e-11, 1.81e-11] | 95% CI [-1.35e-11, 1.00] | 95% CI [-7.05e-11, 4.06e-11] |
|  | Number of Children in Home | W = 2232.5, p-value = 0.80 | W = 2576, p-value = 0.017 | W = 553.5, p-value = 0.30 | W = 599.5, p-value = 0.084 |
|  |  | 95% CI [-1, 1] | 95% CI [ 4.77e-11, 2.00] | 95% CI [-1, 2] | 95% CI [-2.53e-11, 2.00] |
|  | Use a Bank | p-value = 0.017 | p-value = 1 | p-value = 1 | p-value = 0.14 |
|  |  | 95% CI [ 0.072, 0.82] | 95% CI [ 0.29, 2.79] | 95% CI [ 0.12, 4.93] | 95% CI [ 0.042, 1.63] |
| **Fisher's Exact Test for Count Data** | Sex | p-value = 1 | p-value = 0.35 | p-value = 1 | p-value = 1 |
|  |  | 95% CI [ 0.25, 3.48] | 95% CI [ 0.50, 7.42] | 95% CI [ 0.17, 6.39] | 95% CI [ 0.081, 9.10] |
|  | Marital Status | p-value = 0.014 | p-value = 0.64 | p-value = 0.23 | p-value = 0.52 |
|  | Education Level | p-value = 0.11 | p-value = 0.24 | p-value = 0.97 | p-value = 0.25 |
|  | Profession | p-value = 0.26 | p-value = 0.044 | p-value = 0.43 | p-value = 0.26 |
|  | Use a Smartphone to Bank | p-value = 0.23 | p-value = 0.20 | p-value = 0.16 | p-value = 1 |
|  |  | 95% CI [ 0.63, 5.96] | 95% CI [ 0.71,7.16] | 95% CI [ 0.58 ,18.31] | 95% CI [ 0.13, 4.57] |
|  | Subsistence Farmers  Fishers Exact Test for Count Data | p-value = 0.80 | p-value = 0.80 | p-value = 1 | p-value = 0.45 |
|  |  | 95% CI [ 0.28, 2.56] | 95% CI [ 0.26, 2.54] | 95% CI [ 0.16, 4.46] | 95% CI [ 0.072, 2.68] |
|  | Own Land | p-value = 0.89 | p-value = 0.89 | p-value = 1 | p-value = 1 |
|  | Livestock | p-value = 0.18 | p-value = 0.19 | p-value = 1 | p-value = 1 |
|  | Water Source | p-value = 0.53 | p-value = 1 | p-value = 1 | p-value = 1 |
|  |  | 95% CI [ 0.40, 6.42] | 95% CI [ 0.26, 4.54] | 95% CI [ 0.076, 11.10] | 95% CI [ 0.012, 17.45] |
|  | Treating Water | p-value = 0.72 | p-value = 1 | p-value = 0.36 | p-value = 1 |
|  |  | 95% CI [ 0.28, 6.77] | 95% CI [ 0.13, 3.97] | 95% CI [ 0.27, 23.50] | 95% CI [ 0.016, 13.11] |
|  | Water Scarcity | p-value = 1 | p-value = 1 | p-value = 0.48 | p-value = 0.14 |
|  |  | 95% CI [0.29, 2.76] | 95% CI [ 0.28, 2.98] | 95% CI [ 0.32, 10.27] | 95% CI [ 0.57, 22.74] |
|  | Toilet Type | p-value = 1 | p-value = 0.27 | p-value = 1 | p-value = 0.034 |
|  | Toilet Location | p-value = 1 | p-value = 1 | p-value = 1 | p-value = 1 |
|  | Lighting Type | p-value = 0.36 | p-value = 0.28 | p-value = 0.97 | p-value = 0.41 |
|  | Cooking Stove Type | p-value = 0.53 | p-value = 0.72 | p-value = 0.67 | p-value = 0.054 |
|  | Fuel | p-value = 0.85 | p-value = 0.37 | p-value = 0.82 | p-value = 0.037 |
